# Development and Validation of a High Sensitivity Rapid Xylazine Dipstick for Clinical Urine Testing

**DOI:** 10.1101/2025.03.07.25323583

**Authors:** Ping Wang, William Butler, Niluksha Walalawela Abeykoon, Bridgit O. Crews, Xiaofeng Xia

## Abstract

**Background:** Xylazine has been increasingly linked to human overdose deaths. No antidote has been identified and naloxone cannot reverse the effect of xylazine. Xylazine withdrawal is not alleviated by opioids. It is imperative to detect xylazine when treating overdoses. No screening method for xylazine has been approved by FDA. We aim to develop a rapid and high sensitivity xylazine test for clinical urine testing.

**Methods:** Monoclonal antibodies with high sensitivity and specificity against xylazine were developed. The leading clone was used to develop a competitive lateral flow immunoassay. The analytical cutoff, specificity and clinical performance of this test was characterized using standards in drug-free urine and clinical urine samples.

**Results:** The rapid xylazine dipstick test has a test time of 5 minutes, and a cutoff of 10 ng/mL xylazine in drug-free urine. No cross reactivity with other commonly used drugs or endogenous metabolites were observed, except for 3% cross reactivity with clonidine. In 181 mass spectrometry confirmed clinical urine samples with xylazine concentrations ≥10 ng/mL and 120 urine samples with xylazine concentrations <10 ng/mL, the dipstick demonstrated a clinical sensitivity of 100% and a clinical specificity of 97%. All 4 false positives had combined xylazine and 4-hydroxy-xylazine concentrations in the 5-10 ng/mL range, with additional xylazine metabolites detected by mass spectrometry.

**Conclusions:** When used with 10 ng/mL cutoff, the rapid xylazine dipstick demonstrates high clinical sensitivity and clinical specificity in urine samples, compared to gold standard mass spectrometry methods. This novel test has the potential to enable informed clinical decisions in suspected xylazine overdoses.

## Introduction

Xylazine is a non-opioid veterinary tranquilizer and an alpha-2 adrenergic receptor agonist that has been increasingly linked to human overdose deaths in the US. Data from the Drug Enforcement Administration (DEA) indicates xylazine-positive overdose deaths increased by >100-1127% in different US regions from 2020-2021(1). In 2023, fentanyl laced with xylazine was designated as an emerging threat to US public health (2).

Xylazine can slow breathing and heart rate and decrease blood pressure to dangerously low levels. The usual opioid antidote naloxone does not work to reverse the effect of xylazine when given at typical doses and patients may have extended sedation periods after naloxone administration. Xylazine may cause profound mental status depression and serious and extended withdrawal effects that are not alleviated by the administration of opioids and may need to be carefully managed (3). As such, it is imperative to identify xylazine’s presence when treating potential overdoses. Although in certain regions in the US Northeast (eg. Philadelphia), xylazine is present in almost all illegal fentanyl preparations, with median content exceeding fentanyl (4), there are wide geographic variations in xylazine content (5). Besides being mixed with fentanyl, xylazine has also been found in mixtures containing cocaine, heroin and other drugs (1). Therefore, fentanyl testing cannot be reliably used as a substitute for xylazine testing.

There is currently no rapid diagnostic method for xylazine that has been approved or cleared by FDA. A forensic-use only immunoassay for central lab instruments has been reported, using a cutoff of 10 ng/mL (6). Several xylazine testing strips are commercially available for drug-checking purposes but are found to have low sensitivity and poor correlation with mass spectrometry. For example, one commercially available lateral flow immunoassay (LFA) with a cutoff of 500 ng/mL for xylazine was found to have a clinical sensitivity of 74% compared to liquid chromatography tandem mass spectrometry (LC-MS/MS) when evaluated in clinical urine samples (7). Another commercially available LFA with a cutoff of 1000 ng/mL for xylazine in aqueous solution demonstrated 86% clinical sensitivity against mass spectrometry (Unsihuay et al, submitted). Cross reactivity of these assays with lidocaine, levamisole, promethazine, ketamine, methamphetamine and diphenhydramine has been reported (8–13), resulting in false positives when xylazine is not present in urine samples. In clinics and hospitals, overwhelming false positives or false negatives can create workflow and economic challenges, negating the value of rapid screening. In one research study, xylazine haptens were used to generate xylazine-specific mice antibodies using the traditional hybridoma/ascites approach. The LFA developed using such antibodies had a cutoff of 1.6 ng/mL xylazine in spiked human urine diluted by buffer. However, this assay was not tested in real clinical urine samples (14). The requirement of urine dilution before testing is impractical in clinical practice.

In this work, we aim to develop and validate a rapid and sensitive xylazine point of care test for clinical urine samples, with high clinical sensitivity and specificity. The pharmacokinetics and pharmacodynamics of xylazine in humans is poorly understood. A recent study calculated a median terminal half-life for xylazine in individuals who use fentanyl mixed with xylazine of 12 hours (15), suggesting the time window for urine detection may extend several days if methods achieve sufficiently low detection limits. . Current understanding is limited with regards to how xylazine concentrations in blood or urine correlate with pathological effects in humans. In hospitalized overdosed patients testing positive for fentanyl and xylazine, xylazine urine concentrations are found to span a wide range, from 10-20000 ng/mL (6, 16), with a 50^th^ percentile of approximately 1000 ng/mL for xylazine, and >200 ng/mL for 4-OH-xylazine. Median postmortem blood xylazine concentration reportedly range from 16-21 ng/mL in fentanyl-associated fatal overdose cases (17, 18). Initial presenting plasma xylazine concentrations ranged 0.7-127.2 ng/mL in individuals with urine xylazine detected ≥1 ng/mL (15). Based on these datapoints, cutoffs of 500 or 1000 ng/mL for urine screening are likely to miss the majority of cases with xylazine present in the urine. We aim to use 10 ng/mL as the cutoff for urine screening and use the competitive LFA mechanism and a dipstick format to achieve rapid readout.

To achieve this cutoff, one of the most important reagents in the development of this immunoassay is the antibody against xylazine. We used our single B cell technology to rapidly generate and produce highly specific rabbit monoclonal antibodies (mAbs) with sub-nanomolar affinity for xylazine. The mAbs were then first screened using competitive enzyme-linked immunosorbent (ELISA) assays to identify clones with the highest sensitivity and specificity towards xylazine, then screened for suitability for LFA. The leading mAb was used to develop a competitive LFA. Performance of this assay was characterized using xylazine standards in drug-free urine and real-world clinical urine samples.

## Methods

### Materials and Chemicals

KLH and BSA conjugated xylazine were purchased from AAT Bioquest (Pleasanton, CA). Xylazine and Freund’s adjuvants were obtained from Sigma Aldrich (St Louis, MO). Other chemicals for specificity testing were purchased from Sigma Aldrich (St Louis, MO) or Cayman Chemical (Ann Arbor, MI).

### Animal immunization

Rabbit immunization was performed by Cocalico Biologicals (Stevens, PA) following a standard procedure (19). All animal maintenance, care and use procedures were reviewed and approved by the company’s Institutional Animal Care and Use Committee (IACUC). New Zealand White rabbits were immunized subcutaneously with 500 μg of KLH-conjugated xylazine emulsified with complete Freund adjuvant for the first dose and incomplete Freund adjuvant for subsequent boosts. The animals received 2 booster injections at 3-week intervals. The animals were sacrificed for spleen harvest 14 days following the final boost.

### Single B cell antibody cloning

The rabbit spleen was collected for splenocyte harvest. The cells were laid on Lympholyte-Mammal (Cedarlane, Burlington, NC) and centrifuged at 800 g for 30 minutes. The lymphocytes were collected at the interface and washed before stained by biotinylated BSA-xylazine and then AF647-streptavidin (BioLegend, San Diego, CA).

Single antigen-binding B cells were sorted using an S3 cell sorter (Bio-Rad, Hercules, CA), and deposited into a PCR tube with 5 μL lysis buffer (Takara Bio, San Francisco, CA). cDNAs were then prepared by reverse transcriptase (Takara Bio, San Francisco, CA) and the antibody coding regions were amplified by PCR. The PCR products were then cloned into an antibody expression vector with a CMV promoter.

### Recombinant antibody expression

Expi293 cells (Thermofisher, Waltham, MA) were transfected with the antibody expression vectors using Expifectamine (Thermofisher, Waltham, MA) (20). After 5 days the cell culture medium was collected. In screening assays, after 5 days the cell culture medium was collected and used in the immunoassays without purification. For further characterization antibodies were purified by protein A chromatography (Cytiva, Marlborough, MA).

### Antibody screening competitive ELISA assay

To carry out antibody screening, transparent 96-well plates were coated with 1 μg/mL BSA-conjugated xylazine. Cell culture medium was diluted 100 times and added to the wells, with or without 10 ng/mL xylazine, to incubate for 1 hour at room temperature. The wells were washed extensively, and HRP-conjugated goat anti-rabbit secondary antibody (R&D Systems, Minneapolis, MN) was added to incubate for 1 hour. The free xylazine competes with the immobilized BSA-conjugated xylazine, leading to reduced antibody binding to the surface. After washing, TMB substrate (Thermo Fisher, Waltham, MA) was added and the absorbance was read with a plate reader. The xylazine binding affinities of the antibodies were quantified by the inhibition percentage calculated using the following equation:

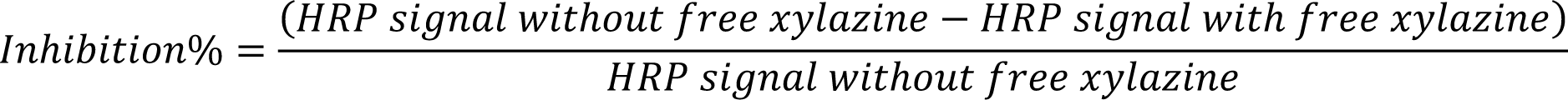

The IC_50_ value was defined as the xylazine concentration that led to 50% inhibition of the ELISA signal compared to the blank control sample.

### Competitive LFA Design

The conjugate pad of the LFA was sprayed with gold nanoparticles conjugated with xylazine-specific mAb. Xylazine-BSA and anti-rabbit Ab are printed onto the test and control lines on the nitrocellulose membrane, respectively.

### LC-MS/MS methods for xylazine quantitation

A subset of 107 urine samples was tested using an LC-MS/MS method developed at Washington University. The details of the method have been published previously for plasma and identical LC-MS/MS parameters were applied to quantify xylazine in urine (15). Urines were prepared using an automated liquid handler (Hamilton Company, Reno, NV) in a 96-well plate. Thirty (30) μL of internal standard solution containing 20 ng/mL xylazine-D6 (Caymen Chemical, Ann Arbor, MI) in 10% methanol and 30 µL of B-one beta-glucuronidase (Kura Biotech, Atlanta, GA) were added to 30 μL of urine specimen, followed by 3 mixing cycles. The plate was held at ambient temperature for 15 minutes, sealed and centrifuged for 5 minutes; 1 μL of prepared specimen was injected for LC-MS/MS analysis. The limit of detection (LOD) of this method was 0.1 ng/mL for xylazine and 0.2 ng/mL for 4-OH-xylazine. The analytical measurement range for quantification was 1-2000 ng/mL for xylazine and 4-OH-xylazine. In addition, xylazine metabolites including oxo-xylazine (m/z 235), OH-oxo-xylazine (m/z 251), sulfone-xylazine (m/z 253) and OH-sulfone-xylazine (m/z 269) were identified qualitatively using product ion scanning with peak areas determined by individual multiple reaction monitoring (MRM) transitions as described previously (14).

A second subset of 121 urine samples was tested using an LC-MS/MS method developed at Jefferson University Hospital. The details of this method have also been published previously (6, 16). The limit of quantitation (LOQ) was 5 ng/mL and the linearity range was 5-30000 ng/mL for xylazine. Urine was tested directly without any pretreatment.

### Gas chromatography mass spectrometry (GC-MS) method for xylazine qualitative detection

A third subset of 71 urine samples was tested using a GC-MS method developed at the Hospital of the University of Pennsylvania. The qualitative GC-MS method screens over 200 substances, including xylazine, confirming presence with a match of retention time and spectra ions with quality score ≥0.8. The limit of detection for substances ranges from 200 to 1000 ng/mL. Only urine samples determined to be positive for xylazine were included in this study.

### Spiked standards testing in drug-free urine

Xylazine, 4-OH-xylazine standards were spiked into certified drug-free urine (UTAK, Valencia, CA) at concentrations of 0, 1, 5 …to 1000 ng/mL and tested using the dipstick following protocol described in Figure 1. For specificity testing, 182 commonly used medications, substances and endogenous metabolites were spiked into drug-free urine at 100 μg/mL or higher and tested using the dipstick. One hundred (100) μL of each sample was used for each test. If cross reactivity was observed at the highest concentration, lower concentrations of the compound standard were tested. Percent cross-reactivity is calculated by dividing the cutoff concentration (10 ng/mL) by the lowest concentration of the compound where the assay response was positive x 100.

**Figure 1.**
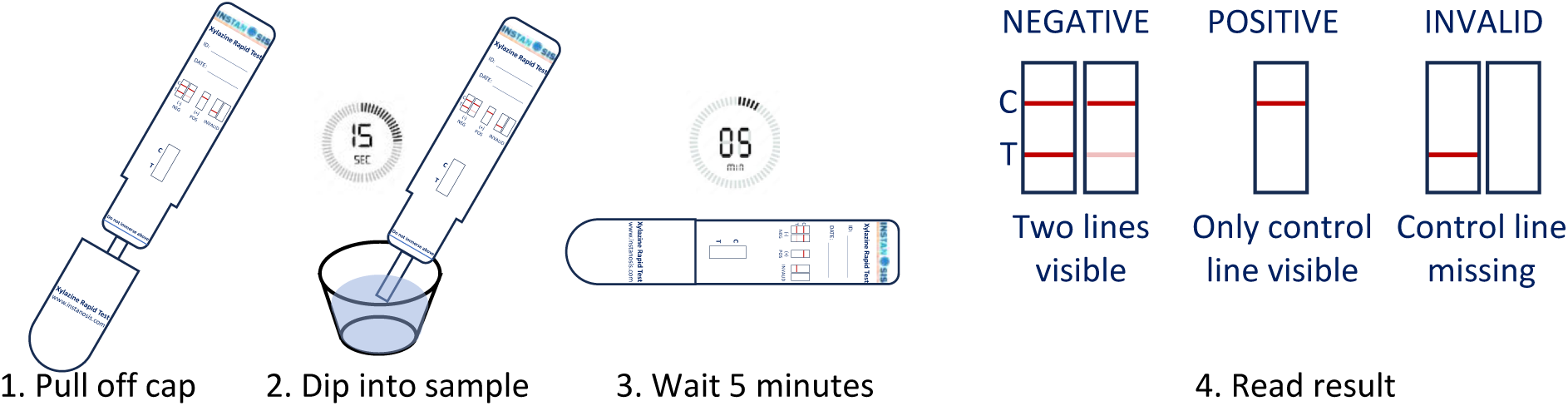
The xylazine dipstick test workflow.

### Clinical sample testing

Deidentified clinical urine samples were collected following protocols approved by the Institutional Review Boards at each institution. One hundred and twenty-one (121) deidentified urine samples were obtained from Jefferson University Hospital. One hundred and seven (107) deidentified urine samples were obtained from Washington University. Seventy-one (71) deidentified urine samples were obtained from the Hospital of the University of Pennsylvania. All samples were stored at -80°C after clinical testing was completed, until the time of analysis using mass spectrometry or dipstick. One hundred (100) μL of each sample was tested following the protocol described in Figure 1, without sample pretreatment. Results were read by three individuals independently. The majority of the readout for each sample was used to calculate clinical sensitivity and specificity.

## Results

### Monoclonal antibodies against xylazine

The results of xylazine mAb developed using the Single B Cell technology are summarized in Figure 2A. Two leading clones I2 and F1, with similar affinity and specificity profiles for xylazine were identified. Both enable cutoffs of ≤ 10 ng/mL in LFA. One of these (I2) was chosen for further LFA development and optimization. The affinities of the mAbs developed in this study for xylazine is characterized in Figure 2B. In comparison, the best commercially available antibody exhibits an IC_50_ of ∼0.35 ng/mL (21). The highest affinity clone developed in this study demonstrated ∼100-fold higher affinity (100-fold lower IC_50_) compared to the commercial antibody.

**Figure 2.**
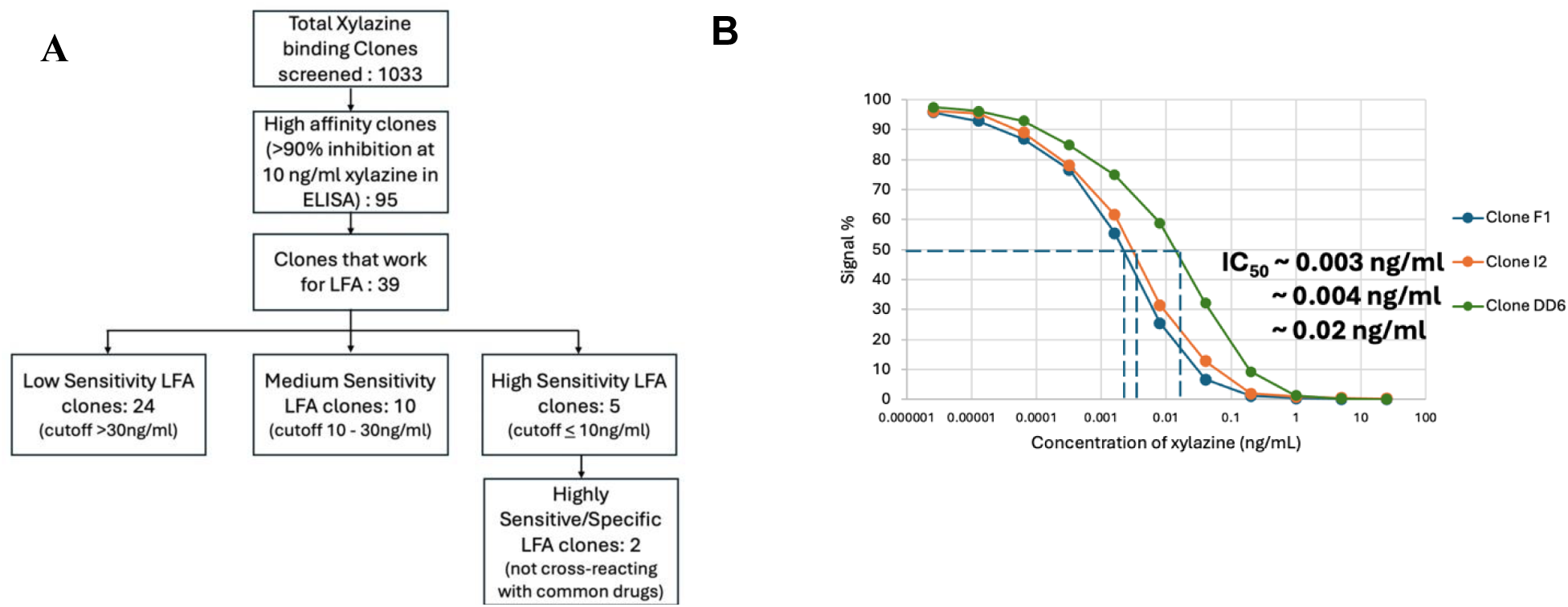
**A**. Flowchart summarizing results of the development of rabbit monoclonal antibodies against xylazine using Single B Cell Express Antibody Discovery (SBEAD). LFA: lateral flow assay. **B**. The antibodies developed in this study exhibit high affinities for xylazine.

### Development and optimization of the xylazine dipstick test

The workflow of the xylazine dipstick test is described in Figure 1. Results are read in 5 minutes. The presence of both control and test lines, including faint test lines, indicate a negative result. The presence of only the control line indicates a positive result.

### Cutoff and specificity/cross-reactivity of the xylazine dipstick test

The cutoff of the xylazine dipstick was evaluated using spiked standards in drug-free urine and compared with 3 commercially available xylazine strip tests. The results are described in Figure 3. All evaluated tests use the competitive immunoassay design. Therefore, the concentration at which the test line first becomes completely invisible (positive test result) as xylazine concentration increases is defined as the cutoff. As indicated by the red arrows, the dipstick test developed in this study had a cutoff of 10 ng/mL xylazine. Two of the other 3 strips had cutoffs of 1000 ng/mL, and cutoff for the last strip was >1000 ng/mL, indicated by the presence of the test line across all tested concentrations.

**Figure 3.**
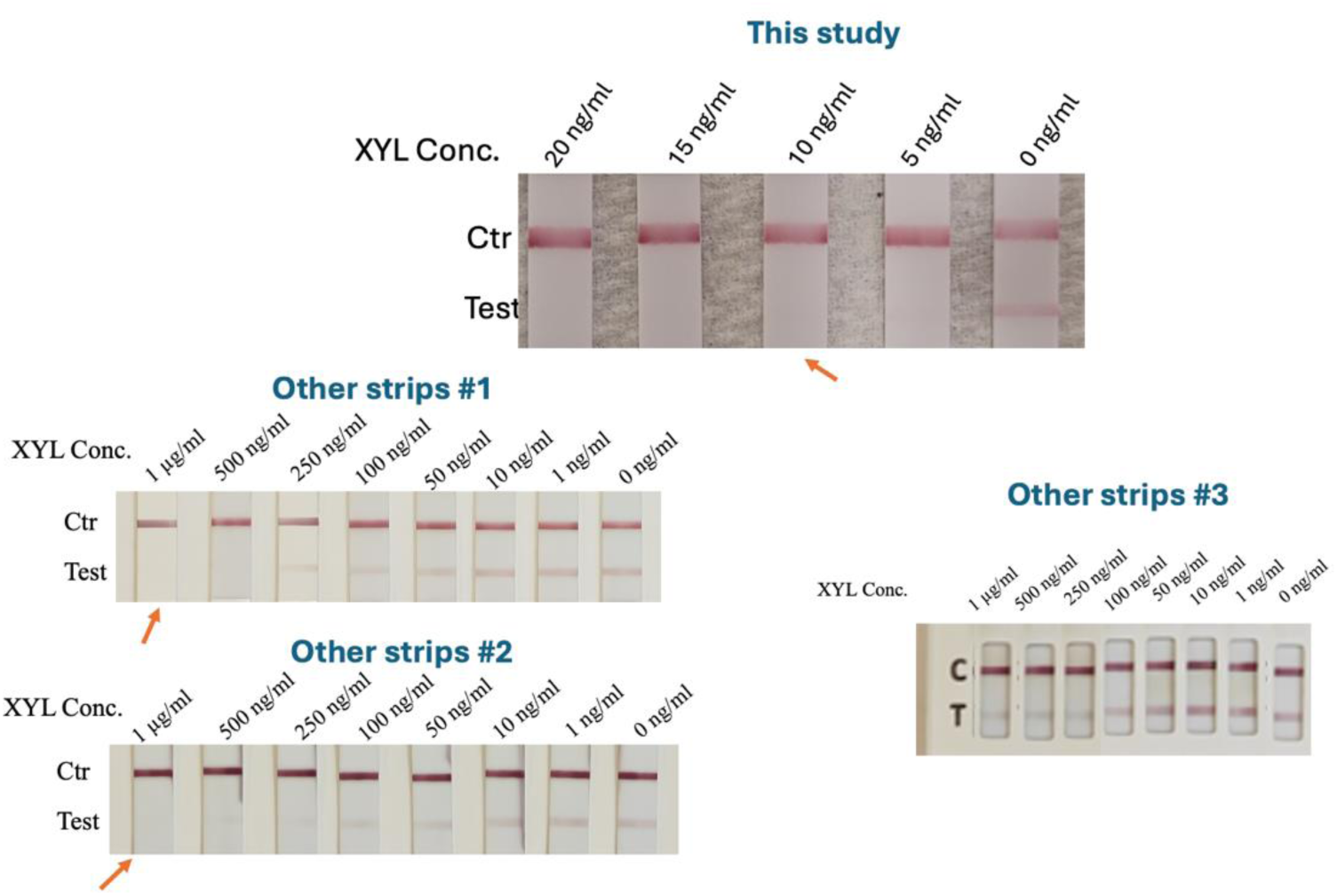
Cutoff characterization for the xylazine dipstick developed in this study and other commercially available strip tests, using standards spiked into drug-free urine. Red arrows point to where cutoffs are (test line first becomes invisible completely as xylazine concentration increases).

The specificity/cross-reactivity of the xylazine dipstick was evaluated by testing 182 commonly used medication, substances and endogenous metabolites spiked into drug-free urine, at the concentration of 100 μg/mL or higher. None of the tested compounds (Supplemental Table 1) demonstrated cross-reactivity with the dipstick, except for clonidine, which had a cross-reactivity of 3% (positive at 300 ng/mL). The xylazine metabolite, 4-OH-xylazine, was tested using the dipstick and found to have 100% cross-reactivity (positive at 10 ng/mL).

### Clinical performance of the xylazine dipstick test

Three sets of deidentified clinical urine samples were collected and tested using the xylazine dipstick test. To assign the gold standard values using mass spectrometry, 2 sample sets were quantitated for xylazine using two LC-MS/MS methods separately (6, 15, 16), with LOQs of 1 and 5 ng/mL xylazine, respectively. One of the LC-MS/MS method also included glucuronide hydrolysis pretreatment, quantitation of 4-OH-xylazine, and identification of other xylazine metabolites including sulfone-xylazine, OH-sulfone xylazine, OH-oxo-xylazine and oxo-xylazine (15). The 3rd sample set was qualitatively tested for xylazine using a GC-MS method. Only xylazine positive urine samples were included in the 3^rd^ set, as the GC-MS cutoff is higher than the dipstick cutoff. Since the dipstick had equal cross-reactivity with xylazine and 4-OH-xylazine, the combined concentration of the two compounds was compared relative to the 10 ng/mL cutoff, where 4-OH-xylazine concentrations were available. The distribution of xylazine concentrations in all urine samples is summarized in Supplemental Table 2.

The dipstick results for all clinical urine samples are summarized in Table 1. Using the 10 ng/mL cutoff, the clinical sensitivity of the dipstick was 100% and clinical specificity was 97%. All 4 false positives had combined xylazine and 4-hydroxy-xylazine concentrations in the 5-10 ng/mL range, with additional xylazine metabolites detected on LC-MS/MS by product ion scanning (Table 2) (15). Sulfone-xylazine and OH-oxo-xylazine were detected in all 4 samples, while OH-sulfone-xylazine was detected in 3 of 4 samples. Due to the lack of commercial standards, we could not determine if these additional metabolites cross react with the dipstick test and contributed to the positive results above the threshold. If 5 ng/mL was used as the cutoff, the same set of clinical data would result in a clinical sensitivity of 98% and a clinical specificity of 100%. Depending on clinical priority, cutoffs between 5-10 ng/mL may be chosen to maximize either clinical sensitivity or clinical specificity.

**Table 1.**
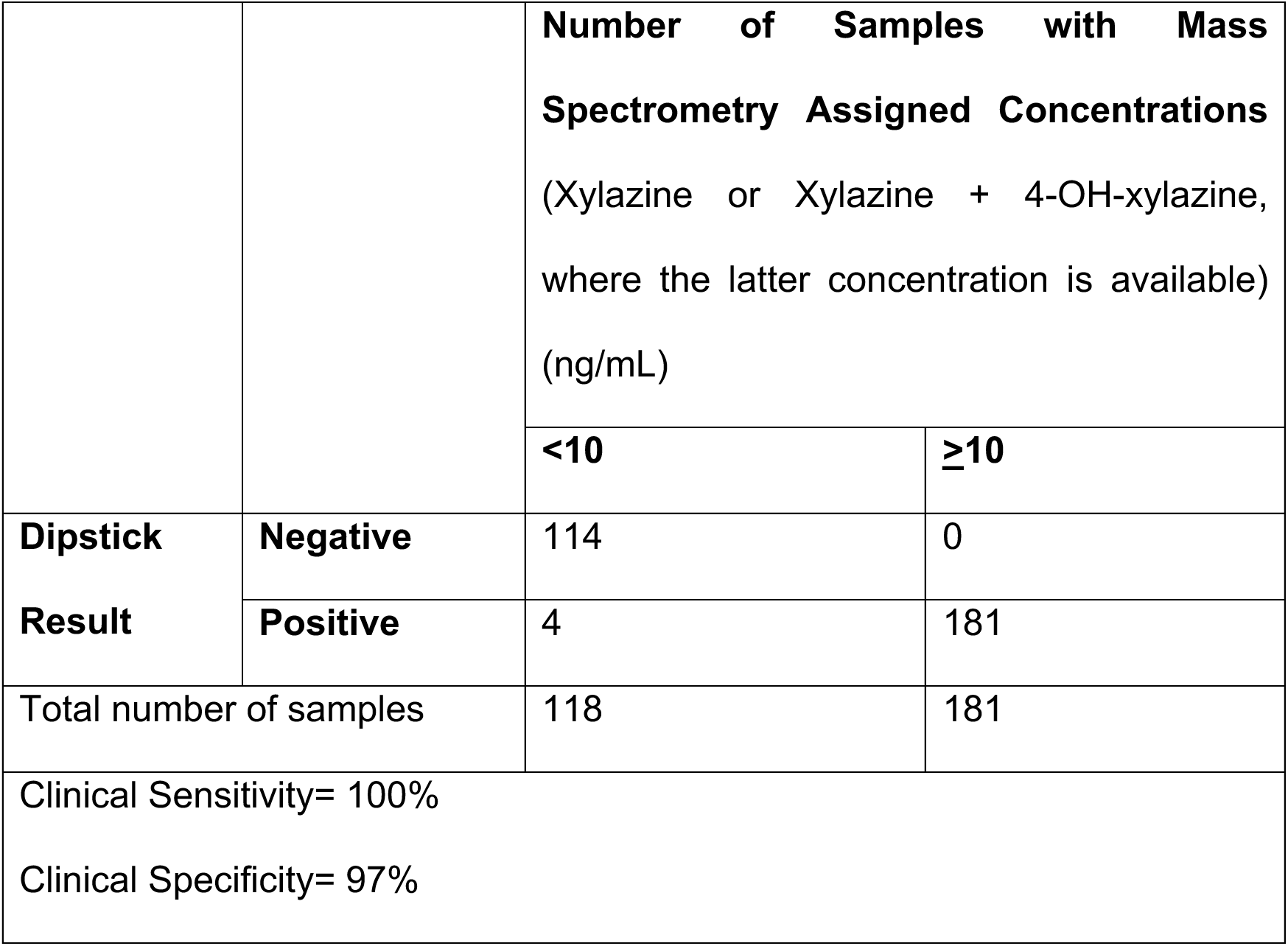
Summary of the xylazine dipstick clinical performance data

|  |  | **Number of Samples with Mass Spectrometry Assigned Concentrations**<br>(Xylazine or Xylazine + 4-OH-xylazine, where the latter concentration is available)<br>(ng/mL) |  |
| --- | --- | --- | --- |
|  |  | **<10** | **≥10** |
| **Dipstick Result** | **Negative** | 114 | 0 |
|  | **Positive** | 4 | 181 |
| Total number of samples |  | 118 | 181 |
| Clinical Sensitivity= 100% |  |  |  |
| Clinical Specificity= 97% |  |  |  |

**Table 2.** Xylazine, 4-OH-xylazine and other metabolites detected on LC-MS/MS of the 4 false positive samples using the 10 ng/mL cutoff.

| Sample ID | Xylazine (ng/mL) | 4-OH-xylazine (ng/mL) | Xylazine+4-OH-xylazine (ng/mL) | Sulfone-xylazine to xylazine peak area ratio | OH-sulfone-xylazine to xylazine peak area ratio | OH-oxo-xylazine to xylazine peak area ratio | Oxo-xylazine to xylazine peak area ratio |
| --- | --- | --- | --- | --- | --- | --- | --- |
| 1 | 3.9 | 1.9 | 5.8 | 6.7 | 0.58 | 4.46 | 0.00 |
| 2 | 6 | 1 | 7 | 1.35 | 0.36 | 1.07 | 0.00 |
| 3 | 6.4 | 2.1 | 8.5 | 3.54 | 0.80 | 2.72 | 0.00 |
| 4 | 6.7 | 0.6 | 7.3 | 0.86 | 0.00 | 0.57 | 0.00 |

## Discussion

In this study we first developed mAbs with high affinity and specificity towards xylazine. A rapid xylazine dipstick test was subsequently developed and validated with a cutoff of 10 ng/mL, ∼100 fold higher analytical sensitivity than commercially available xylazine strip tests, and high clinical sensitivity and specificity. As far as we know, this is the first time a point-of-care xylazine screening test with such high sensitivity has been reported. This higher sensitivity will allow identification of most patients exposed to xylazine. The high specificity means high confirmation rate by mass spectrometry when screen-positive samples are sent to confirmation. This can help build trust for the rapid dipstick test, address workflow and economic concerns created by high number of false positives and boost clinical adoption. With the 5-minute test time and easy-to-use format, this novel test has the potential to enable quick informed clinical decisions in cases of suspected xylazine overdose.

MAbs with different tiers of affinities for xylazine were developed during this process. Although we only focused on LFA development in this study, these antibodies may be leveraged to develop other novel diagnostics with different sensitivities or cutoffs for xylazine.

The choice of the 10 ng/mL cutoff is relatively arbitrary, since it is currently unclear what level of xylazine urine concentration is considered clinically important. However, the excellent correlation between the dipstick and the LC-MSMS methods, which have lower limits of detection and will serve as confirmation methods in clinical testing, suggests this cutoff is reasonable.

One limitation of the study is that different mass spectrometry methods were used to assign ground truth values. The two LC-MS/MS methods have slightly different LOQs, although both are lower than the dipstick cutoff. It is unknown how the two LC-MS/MS methods correlate with each other. Glucuronide hydrolysis pretreatment was conducted for only one of the LC-MS/MS methods, potentially resulting in higher overall xylazine and 4-OH-xylazine concentrations. It is unknown if xylazine-glucuronide or 4-OH-xylazine-glucuronide are major metabolites excreted in urine or whether they would cross react with the dipstick test. In addition, 4-OH-xylazine quantitation and the detection of other xylazine metabolites was not available for all the samples. Other metabolites may also be present that could cross react. This represents the current real-world situation, as most clinical laboratories only measure/detect the parent drug xylazine if such methods are offered at all.

Due to the lack of commercial standards, the cross reactivity of the dipstick test with other xylazine metabolites (sulfone-xylazine, OH-sulfone xylazine, OH-oxo-xylazine, oxo-xylazine and their glucuronide conjugates) observed on the LC-MS/MS could not be assessed. It is possible that such cross reactivity may have contributed to the positive test signals in the dipstick test. From the perspective of assessing xylazine exposure, all 4 “false positive” samples had definitive evidence of xylazine present at concentrations near the cutoff with additional metabolites detected and therefore may be considered “true positives” in terms of xylazine exposure.

The dipstick test had 3% cross-reactivity with clonidine. Clonidine is also an alpha-2 adrenergic receptor agonist, with a chemical structure highly similar to xylazine.

Although clonidine is often used to manage opioid withdrawal symptoms, blood concentrations are typically only single digit ng/mL in clonidine overdose cases. Obtundedness occurs at blood clonidine concentration <100 ng/mL (22, 23). Although there is no published data on corresponding urine clonidine concentrations in such cases, we believe false positivity due to urine clonidine cross reacting at or above 300 ng/mL would be extremely rare in real world clinical cases.

## Funding/Conflict of Interest

The study was funded by a National Institute on Drug Abuse grant 1U44DA060264 to Ping Wang and Xiaofeng Xia. The other authors have no conflict of interest.

## Data Availability

All data produced in the present work are contained in the manuscript

## Acknowledgements

We thank Dr. Douglas Stickle at Jefferson University Hospital for providing some de-identified urine samples and LC-MS/MS analysis of these samples.

---

**Supplemental Table 1.**
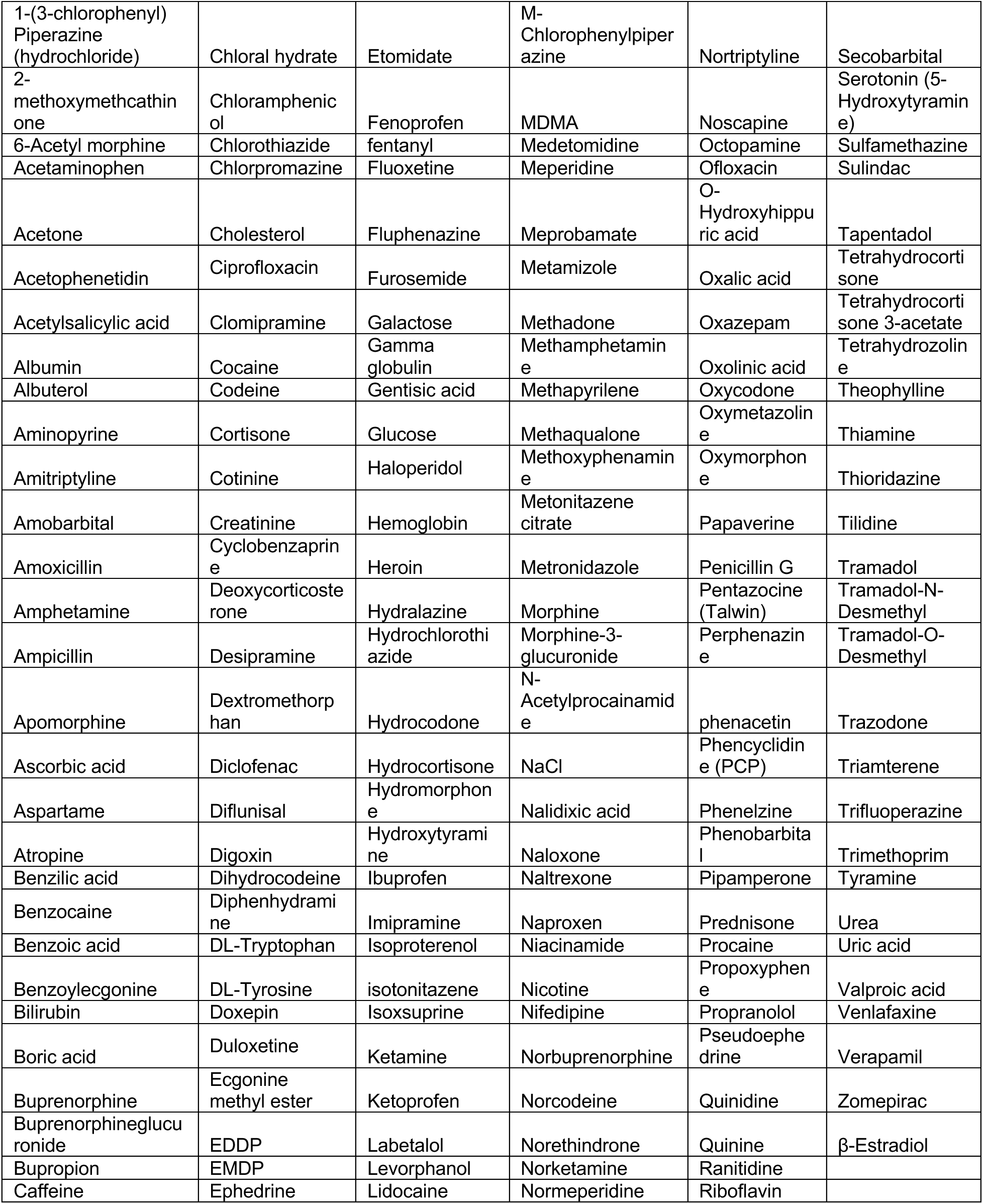
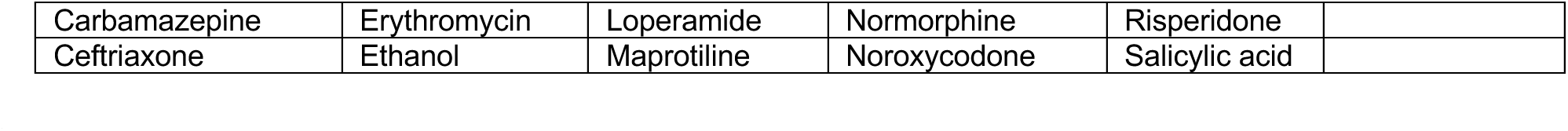
Compound tested with no cross-reactivity with the xylazine dipstick test.

**Supplemental Table 2.**
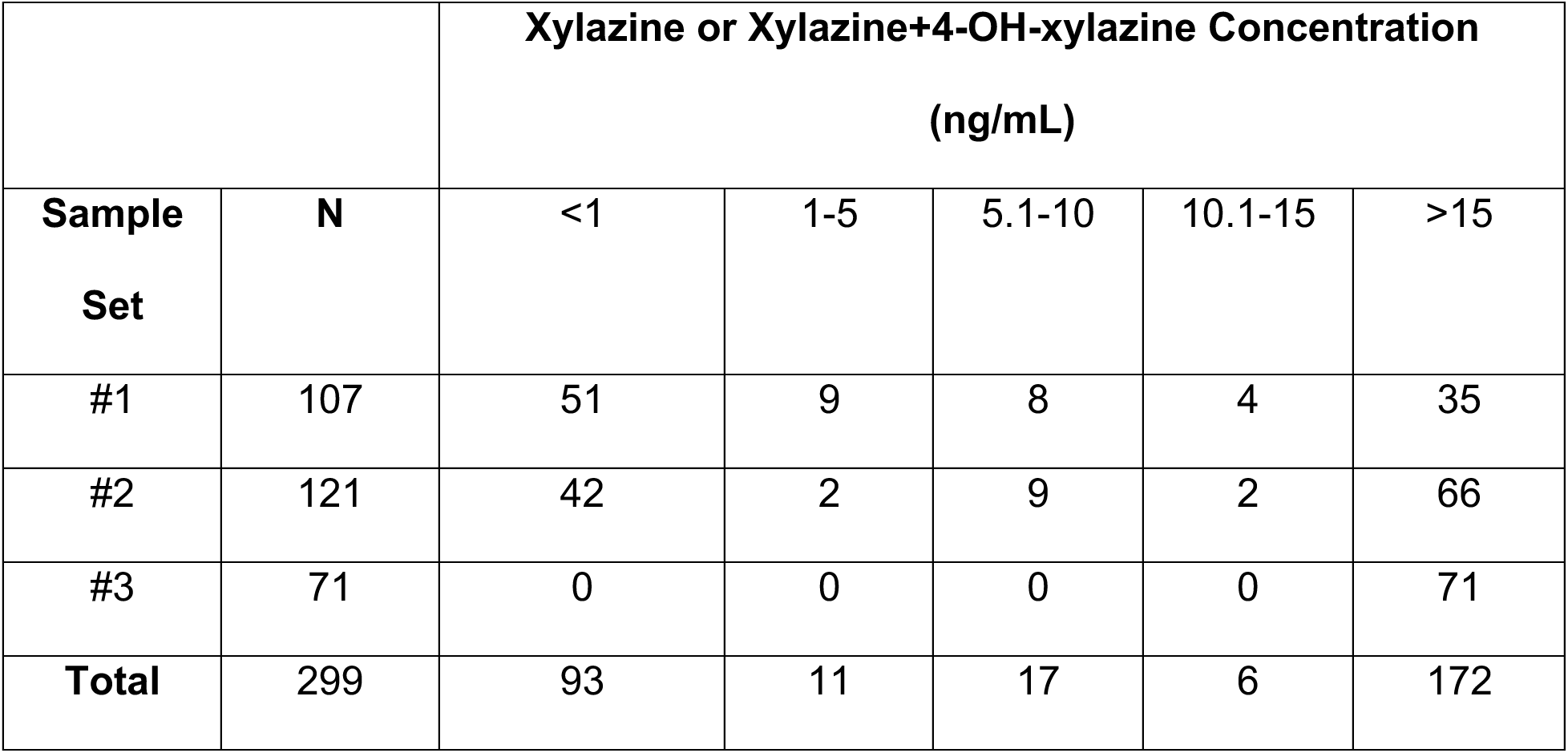
Distribution of xylazine concentrations in clinical urine samples

|  |  | **Xylazine or Xylazine+4-OH-xylazine Concentration** |  |  |  |  |
| --- | --- | --- | --- | --- | --- | --- |
|  |  | **(ng/mL)** |  |  |  |  |
| **Sample Set** | **N** | <1 | 1-5 | 5.1-10 | 10.1-15 | >15 |
| #1 | 107 | 51 | 9 | 8 | 4 | 35 |
| #2 | 121 | 42 | 2 | 9 | 2 | 66 |
| #3 | 71 | 0 | 0 | 0 | 0 | 71 |
| **Total** | **299** | **93** | **11** | **17** | **6** | **172** |

## Notes

### Author Declarations

IRB of University of Pennsylvania gave ethical approval for this work. IRB of Washington University gave ethical approval for this work.

## References

1. Administration UDoJDE. The Growing Threat of Xylazine and its Mixture with Illicit Drugs. October 2022.

2. Biden-Harris Administration Designates Fentanyl Combined with Xylazine as an Emerging Threat to the United States. April 12, 2023.

3. Robert D. Glatter MJLDO, MD. . What’s Best for Treating Patients Who Are on Xylazine or “Tranq”? 2022.

4. DeBord J SJ, Russell R, Denn M, Quinter A, Logan BK, Teixeira da Silva D, and Krotulski AJ. Center for Forensic Science Research and Education. Drug Checking— Quarterly Report. 2024.

5. Friedman J, Montero F, Bourgois P, Wahbi R, Dye D, Goodman-Meza D, Shover C. Xylazine spreads across the US: A growing component of the increasingly synthetic and polysubstance overdose crisis. Drug Alcohol Depend 2022 Apr 1;233:109380. Epub 20220226 as doi: 10.1016/j.drugalcdep.2022.109380.

6. Kyle PB, Mattiello CJ, Hua A, Toohey JM, Korn WR, Saldana-Reed A, Stickle DF. Evaluation of the ARK Diagnostics immunoassay for qualitative detection of xylazine in urine. J Anal Toxicol 2024 Jul 13;48 6:468–71 as doi: 10.1093/jat/bkae043.

7. Contella L, Snyder ML, Kang P, Tolan NV, Melanson SEF. Clinical performance of a new lateral flow immunoassay for xylazine detection. Clin Chem Lab Med 2024 Oct 14. Epub 20241014 as doi: 10.1515/cclm-2024-0947.

8. Krotulski AJ SJ, Debord J, Teixeira da Silva D, Logan BK. Evaluation of Xylazine Test Strips (BTNX) for Drug Checking Purposes, Center for Forensic Science Research and Education, United States. 2023.

9. Sisco E, Nestadt DF, Bloom MB, Schneider KE, Elkasabany RA, Rouhani S, Sherman SG. Understanding sensitivity and cross-reactivity of xylazine lateral flow immunoassay test strips for drug checking applications. Drug Test Anal 2024 Sep;16 9:942–7. Epub 20231203 as doi: 10.1002/dta.3612.

10. M. L. Testing the Tests. 2023.

11. Thompson E, Tardif J, Ujeneza M, Badea A, Green TC, McKee H, McKenzie M, et al. Pilot findings on the real-world performance of xylazine test strips for drug residue testing and the importance of secondary testing methods. Drug Alcohol Depend Rep 2024 Jun;11:100241. Epub 20240506 as doi: 10.1016/j.dadr.2024.100241.

12. Scott L DK, Park JN, Majeed S. Evaluating the Sensitivity, Selectivity, and Cross-reactivity of Lateral Flow Immunoassay Xylazine Test Strips. ChemRxiv.

13. Friedman JR, Montoya AG, Ruiz C, Tejeda MAG, Segovia LA, Godvin ME, Sisco E, et al. The Detection of Xylazine in Tijuana, Mexico: Triangulating Drug Checking and Clinical Urine Testing Data. medRxiv 2024 Aug 22. Epub 20240822 as doi: 10.1101/2024.08.19.24312273.

14. Luo L, Zhang Y, Zhao X, Wu W, Fei J, Yu X, Wen K, et al. Rational hapten design, antibody preparation, and immunoassay development for rapid screening xylazine in biological samples. Food Chem 2025 Feb 15;465 Pt 2:142054. Epub 20241117 as doi: 10.1016/j.foodchem.2024.142054.

15. Lin Y, Farnsworth CW, Azimi V, Liss DB, Mullins ME, Crews BO. Xylazine Pharmacokinetics in Patients Testing Positive for Fentanyl and Xylazine. Clin Chem 2025 Feb 3;71 2:266–73 as doi: 10.1093/clinchem/hvae163.

16. Korn WR, Stone MD, Haviland KL, Toohey JM, Stickle DF. High prevalence of xylazine among fentanyl screen-positive urines from hospitalized patients, Philadelphia, 2021. Clin Chim Acta 2021 Oct;521:151–4. Epub 20210712 as doi: 10.1016/j.cca.2021.07.010.

17. Hays HL, Spiller HA, DeRienz RT, Rine NI, Guo HT, Seidenfeld M, Michaels NL, et al. Evaluation of the relationship of xylazine and fentanyl blood concentrations among fentanyl-associated fatalities. Clin Toxicol (Phila) 2024 Jan;62 1:26–31. Epub 20240214 as doi: 10.1080/15563650.2024.2309326.

18. Korona-Bailey J, Onyango E, Hall KF, Jayasundara J, Mukhopadhyay S. Xylazine-Involved Fatal and Nonfatal Drug Overdoses in Tennessee From 2019 to 2022. JAMA Netw Open 2023 Jul 3;6 7:e2324001. Epub 20230703 as doi: 10.1001/jamanetworkopen.2023.24001.

19. Starkie DO, Compson JE, Rapecki S, Lightwood DJ. Generation of Recombinant Monoclonal Antibodies from Immunised Mice and Rabbits via Flow Cytometry and Sorting of Antigen-Specific IgG+ Memory B Cells. PloS one 2016;11 3:e0152282 as doi: 10.1371/journal.pone.0152282.

20. Xu X, Gao D, Wang P, Chen J, Ruan J, Xu J, Xia X. Epicient homology-directed gene editing by CRISPR/Cas9 in human stem and primary cells using tube electroporation. Scientific reports 2018 Aug 3;8 1:11649 as doi: 10.1038/s41598-018-30227-w.

21. Diagnostics C. Xylazine Antibodies & Antigens.

22. Romano MJ, Dinh A. A 1000-fold overdose of clonidine caused by a compounding error in a 5-year-old child with attention-deficit/hyperactivity disorder. Pediatrics 2001 Aug;108 2:471–2 as doi: 10.1542/peds.108.2.471.

23. RC B. Disposition of Toxic Drugs and Chemicals in Man. 2008.

